# Towards Causal Interpretability in Deep Learning for Parkinson’s Detection from Voice Data

**DOI:** 10.1101/2025.04.25.25326311

**Authors:** Aniruth Ananthanarayanan, Sudeep Senivarapu, Anishsairam Murari

**Author notes:** Authors contributed equally to this work.

## Abstract

This research introduces a comprehensive framework for Parkinson’s Disease (PD) detection using voice recording data. We implemented and evaluated multiple deep learning models, including a baseline Convolutional Neural Network (CNN), an uncertainty-aware Monte Carlo-Dropout CNN (MCD-CNN), as well as a few-shot learning approach to address dataset size limitations. Our models achieved an accuracy over 90% in classifying PD patients using vocal biomarkers, with the ensemble model demonstrating the highest performance. We employed data augmentation techniques to address class imbalance and enhance generalization. Causal feature analysis revealed that the Noise-to-Harmonics Ratio (NHR), Recurrence Period Density Entropy (RPDE), and MDVP jitter parameters were among the most significant vocal biomarkers for PD detection, in order of estimated effect magnitude. Across deep learning models, features exhibiting the strongest absolute correlation with outputs consistently showed the largest estimated effect magnitudes. The few-shot learning approach showed promising results as well, even with limited training examples. This work demonstrates the use of causal feature analysis to validate the analysis of deep learning models, potentially enabling accessible and interpretable non-invasive screening tools.

## Introduction

Parkinson’s disease (PD) is a progressive neurodegenerative disorder that affects more than 10 million people worldwide and can impose significant clinical and economic burdens [1, 2]. Clinical diagnosis typically relies on motor symptoms such as tremor, bradykinesia, rigidity, and postural instability that emerge only after substantial dopaminergic neuron loss [3, 4]. Although modalities such as DaT-SPECT imaging, wearable movement sensors, smartphone-based digital biomarkers, EEG screening, and blood-based molecular assays offer noninvasive or minimally invasive early detection options, they depend on handcrafted features, lack uncertainty quantification, and do not establish causal links between biomarkers and predictions [5–9]. However, hypokinetic dysarthria often manifests up to five years before motor signs, positioning voice recordings as a low-cost and widely accessible screening avenue [4].

This research introduces a comprehensive framework for PD detection from sustained vowel recordings by implementing and evaluating three deep learning models: a baseline Convolutional Neural Network (CNN), an uncertainty-aware Monte Carlo-Dropout CNN (MCD-CNN), and a few-shot learning variant to mitigate the limited amount of data [10, 11]. Our model surpasses 90% accuracy in distinguishing PD from healthy voices, leveraging data augmentation to correct class imbalance and improve generalization. Critically, we apply causal feature analysis to quantify the effect sizes of vocal biomarkers, identifying Noise-to-Harmonics Ratio (NHR), Recurrence Period Density Entropy (RPDE), and MDVP jitter as the most influential predictors [12]. Features exhibiting the highest absolute correlations also demonstrate the largest estimated causal effects across models, confirming their mechanistic relevance.

Shallow classifiers using handcrafted acoustic features (jitter, shimmer, MFCCs) have achieved up to 95% accuracy in voice-based PD detection but remain correlational and opaque [13]. Wearable inertial sensors and gait-analysis platforms reliably quantify gait abnormalities for prodromal motor anomalies [6, 14], while smartphone and smartwatch digital biomarker systems facilitate continuous remote monitoring of both motor and non-motor symptoms [7]. EEG-based screening demonstrates high specificity in early PD neural signatures [8], and blood-based tRNA fragment assays yield AUCs (area under the receiver operating characteristic curve, a measure of classification performance where 1.0 indicates perfect accuracy) around 0.86 for molecular detection [9]. None of these approaches, however, couple deep learning performance with rigorous uncertainty quantification and causal validation of feature importance, which are gaps the new framework addresses.

## Materials and Methods

### Dataset

This study utilizes the Parkinson’s dataset from the UCI Machine Learning Repository [15, 16]. The dataset contains 195 voice measurements from 31 individuals, 8 of whom are healthy and 23 diagnosed with Parkinson’s disease. These voice recordings are characterized by a range of vocal features such as fundamental frequency (jitter), amplitude variation (shimmer), noise-to-harmonics ratio (NHR), and other signal processing metrics commonly known to be indicative of Parkinsonian speech impairments. Each set of signal processing metrics for each voice sample is associated with a label that indicates the individual’s Parkinson’s diagnosis, where 0 represents healthy and 1 represents a patient diagnosed with Parkinson’s. To enhance the quality and diversity of the training data, a range of data augmentation techniques was applied with the goal of balancing class distributions and expanding the dataset. These techniques included adding Gaussian noise to simulate measurement variability, perturbing a subset of features to mimic natural fluctuations, interpolating between samples of the same class to generate intermediate examples, applying global scaling and shifting to introduce broader variability, and randomly masking features to simulate missing or occluded data. Together, these strategies produced a more balanced and representative dataset that can help improve model generalization and reduce bias toward overrepresented classes.

**Figure 1.**
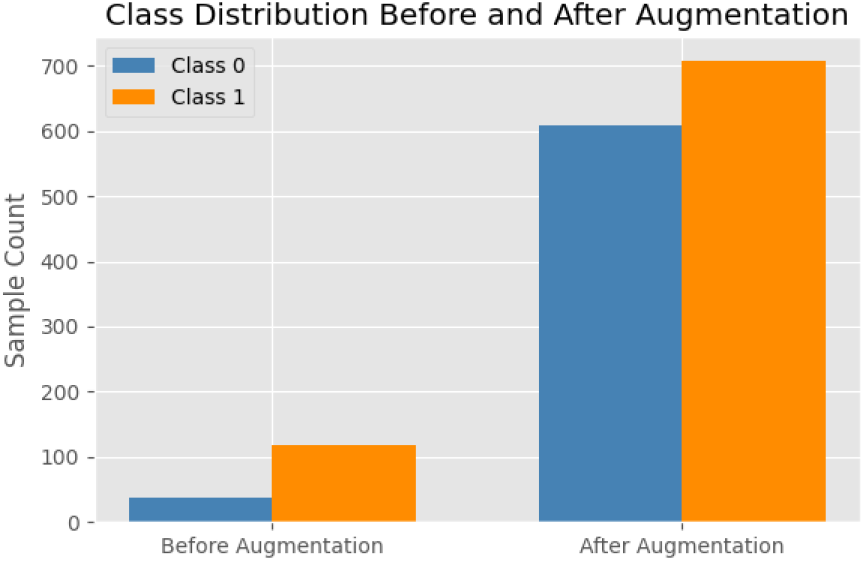
Visualization of class imbalance in UCI ML Repository Parkinson’s dataset. We performed data augmentation to address class imbalance and limited dataset size. However, we did not even out the classes completely as it would have limited the maximum dataset size without diluting the density of the unaugmented samples. Our initial class ratio was about 26:74, and after augmentation, it improved to approximately 46:54.

### Model Architectures

We implemented four complementary deep learning models for Parkinson’s Disease (PD) classification based on acoustic features:

1. **Vanilla CNN**. A baseline 1D convolutional neural network that treats each of the 22 acoustic features as a discrete signal channel. The input is reshaped to (1 *× D*) and passed through two convolutional layers with 4 and 8 filters respectively (kernel size = 3, padding = 1), each followed by ReLU activation and 20% dropout. The output is flattened and passed through a fully connected layer (16 units, ReLU, 20% dropout) before reaching a final sigmoid output neuron for binary classification.
2. **MC-Dropout CNN**. Extends the Vanilla CNN by enabling dropout at inference time via Monte Carlo Dropout. At test time, 50 stochastic forward passes are performed to obtain a distribution over predictions. The final prediction is the mean of these outputs, and model uncertainty is quantified via the predictive variance (epistemic uncertainty).
3. **Few-Shot Learner**. Augments the base CNN with an 8-dimensional prototype embedding layer inserted after the convolutional blocks. During training, embeddings from a small support set (*k* examples per class) are extracted and stored. At inference, query embeddings are compared to support embeddings via Euclidean distance, and predictions are computed as a weighted average based on similarity. This prototypical approach is robust to limited labeled data and encourages generalization from few examples.
4. **Ensemble Model**. Combines the strengths of the Vanilla CNN and MC-Dropout CNN by averaging their output probabilities. This ensemble approach reduces variance and enhances predictive stability by blending deterministic and uncertainty-aware inference.

### Training

All models were trained under consistent optimization settings to ensure fair comparison. We used binary cross-entropy loss and the Adam optimizer with an initial learning rate of 1*×*10^*−*3^. A `ReduceLROnPlateau` scheduler (factor = 0.5, patience = 5) was employed to adapt the learning rate based on stagnation in validation loss. Early stopping with a patience of 10 epochs was applied, restoring the best-performing model weights.

Training was performed with a batch size of 32 for up to 50 epochs. At each epoch, we tracked training/validation loss, validation accuracy, and AUC-ROC. Learning rate scheduling and early stopping jointly contributed to stable convergence and reduced overfitting.

The dataset was split into 72% training, 8% validation, and 20% test data. Validation data was sampled from 10% of the original training split. Model parameters were optimized using the training set, hyperparameters were tuned via validation, and final performance was evaluated solely on the held-out test set to ensure unbiased assessment.

### Causal Analysis

To rigorously assess the extent to which our deep learning models rely on mechanistic biomarkers rather than spurious correlations, we adopt a double/debiased machine learning (DML) framework using the CausalForestDML estimator from the EconML library [17]. Let *Y ∈ {*0, 1*}* denote the binary PD status and let *X* = (*X*_1_, … , *X*_*p*_) be the vector of vocal features. For each feature *X*_*j*_, we define the potential outcome *Y*_*i*_(*x*_*j*_) as the PD probability under an individual-level intervention that sets *X*_*j*_ to value *x*_*j*_ while holding all other covariates fixed. Under the **unconfoundedness** assumption

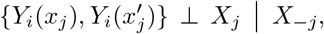

and the **overlap** condition Pr(*ϵ < f*_*j*_(*X*_*−j*_) *<* 1 − *ϵ*) = 1 for some *ϵ >* 0, the conditional average treatment effect (CATE)

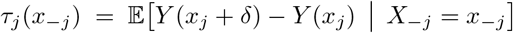

is identified. The CausalForestDML procedure proceeds as follows:

1. **Nuisance estimation** Estimate the outcome regression *m*(*X*_*−j*_) = 𝔼[*Y* | *X*_*−j*_] and the propensity function *g*_*j*_(*X*_*−j*_) = 𝔼[*X*_*j*_ | *X*_*−j*_] via random forest regressors using cross-fitting.
2. **Orthogonalization** For each observation *i*, compute the orthogonalized score

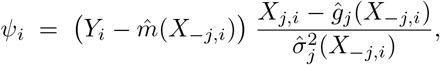

which yields a root-*n*-consistent estimate of *τ*_*j*_.
3. **Aggregation** Obtain individual CATE estimates 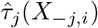 and average over all *n* samples to compute the overall average treatment effect (ATE):

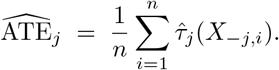
4. **Group-level effects** To evaluate biomarker categories (e.g. MDVP features), we compute the mean of 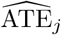 across features within each group.

This DML-based approach accounts for confounding among vocal features, delivers causal effect estimates with desirable statistical properties, and provides interpretable effect-size measures for each vocal biomarker.

## Results

The Vanilla CNN achieved the highest raw accuracy (97.4%) and AUC-ROC (0.9897), while the MC-Dropout CNN provided uncertainty-aware predictions with slightly lower accuracy (89.7%, AUC-ROC = 0.9655). The ensemble balanced these, yielding 92.3% accuracy and AUC-ROC = 0.9862.

**Table 1.**
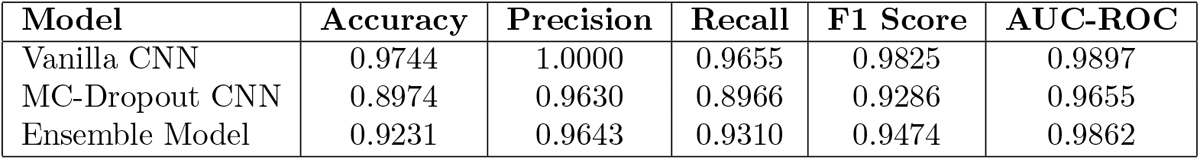
Model comparison. Performance comparison of Vanilla CNN, MC-Dropout CNN, and Ensemble Model on training dataset.

| Model | Accuracy | Precision | Recall | F1 Score | AUC-ROC |
| --- | --- | --- | --- | --- | --- |
| Vanilla CNN | 0.9744 | 1.0000 | 0.9655 | 0.9825 | 0.9897 |
| MC-Dropout CNN | 0.8974 | 0.9630 | 0.8966 | 0.9286 | 0.9655 |
| Ensemble Model | 0.9231 | 0.9643 | 0.9310 | 0.9474 | 0.9862 |

Causal feature ranking identified Noise-to-Harmonics Ratio (NHR), Recurrence Period Density Entropy (RPDE), and MDVP jitter as the top three drivers of model predictions, consistent with their high correlation and large estimated causal effects.

The few-shot learner, even with as few as three examples per class (which sums up to a total of six labeled examples), attained mean accuracy ≈66% and F1 *>* 0.7 on 1,000 runs, outperforming a random forest baseline demonstrating robustness under extreme data scarcity.

**Figure 2.**
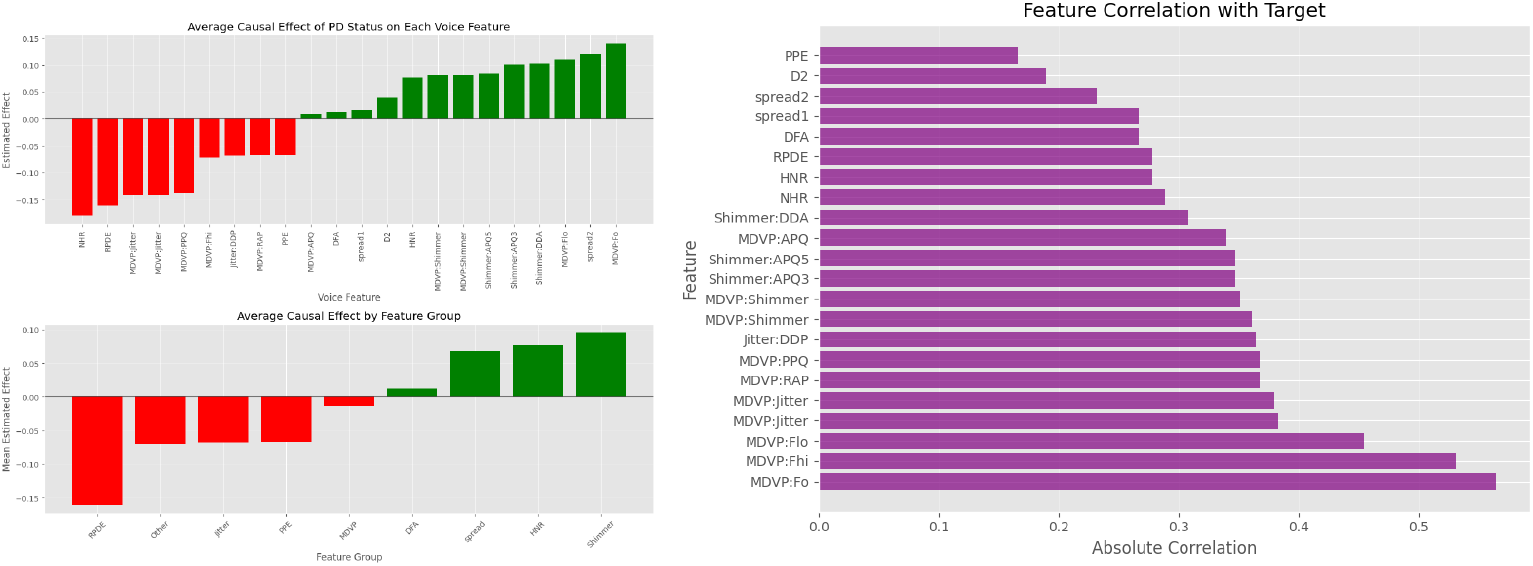
Causation and Correlation. Left: Visualization of estimated effect of various biomarkers and biomarker groups quantified through the CausalForestDML approach. Right: Feature correlation values for Vanilla CNN.

**Figure 3.**
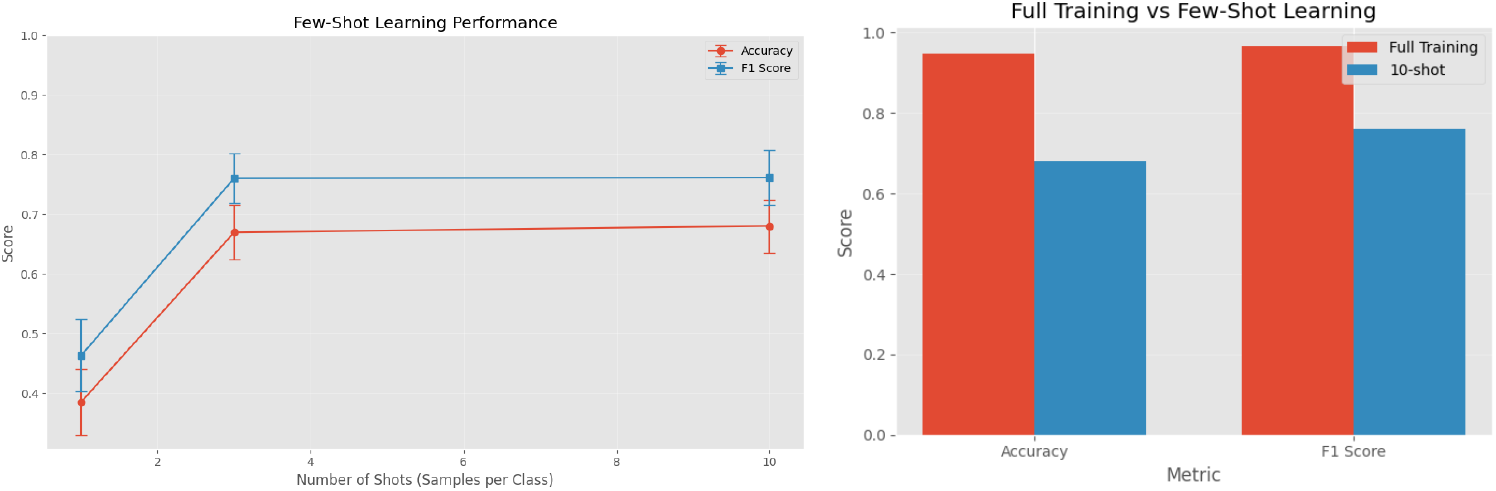
Few-shot learning performance and comparison to full-data training. Left: Mean accuracy (red) and F1 score (blue) ±1 SD of the prototypical few-shot learner as a function of the number of support examples per class. Right: Bar chart contrasting the full-data CNN against the 10-shot few-shot model on test accuracy and F1 score.

## Discussion

This study demonstrates the viability of deep learning for non-invasive, voice-based Parkinson’s disease (PD) screening, achieving both high classification accuracy and interpretable insight into underlying vocal biomarkers. Among the evaluated models, the Vanilla CNN achieved state-of-the-art performance (accuracy: 97.4%, AUC-ROC: 0.9897), outperforming traditional shallow classifiers based on hand-engineered features [13]. While the MC-Dropout CNN exhibited slightly lower predictive accuracy (89.7%, AUC-ROC: 0.9655), it offered a critical advantage: the ability to estimate epistemic uncertainty through stochastic inference, a requirement for clinical decision-making pipelines where confidence calibration can guide further testing or referral [10].

Interpretability was further advanced through causal analysis using the CausalForestDML estimator. This approach identified the Noise-to-Harmonics Ratio (NHR), Recurrence Period Density Entropy (RPDE), and MDVP jitter as features with the highest estimated average treatment effects (ATE) on PD classification probability. These findings align with clinical understanding: jitter captures irregular vocal fold vibrations, NHR reflects inefficient glottal closure, and RPDE quantifies nonlinear voice signal patterns [18, 19]. All are linked to well-established PD symptoms such as dysphonia, bradykinesia, and disrupted motor coordination [20–22]. By combining correlation-based model saliency with causal effect estimation, we show that these features are not only predictive but plausibly mechanistic, enhancing trust in the model’s outputs beyond black-box associations.

The few-shot learner achieved encouraging results in low-data regimes, attaining over 66% accuracy and an F1 score above 0.7 with only three labeled examples per class. This suggests strong potential for deployment in resource-limited settings or early-stage studies, where collecting large-scale labeled voice corpora is infeasible. Prototypical embeddings provided an interpretable basis for class decision boundaries while requiring minimal supervision, demonstrating the model’s promise for scalable clinical screening under data scarcity.

Despite these advances, several limitations warrant attention. First, all models were trained on the UCI Parkinson’s dataset, which includes only 31 unique subjects [15, 16]. While data augmentation increased apparent sample diversity, synthetic perturbations may not capture the full variability of real-world voice recordings across demographics, accents, and environmental conditions. Second, the causal inference methods assume no unobserved confounding; although key features were included, future work should incorporate demographic, comorbidity, and recording-device covariates to further strengthen causal validity [17]. Finally, while the MC-Dropout model estimates epistemic uncertainty, it does not account for aleatoric noise, which could be addressed through heteroscedastic modeling.

Looking forward, expanding this framework to large-scale, longitudinal datasets collected from heterogeneous sources is essential for evaluating generalizability and robustness. Integration with mobile platforms and telehealth systems could enable real-time, non-invasive PD screening in home or rural environments. Here, uncertainty estimates could assist triage workflows, flagging high-risk individuals for follow-up.

Further, combining voice-based models with other digital phenotyping modalities—such as gait data [6, 14], smartphone-based behavior tracking [7], EEG biomarkers [8], or molecular diagnostics [9]—could enhance early detection, provide cross-modal redundancy, and uncover novel PD subtypes.

Notably, current clinical diagnosis of PD largely depends on motor symptom evaluation, with vocal symptoms like dysphonia underutilized despite their diagnostic potential. Our findings suggest that voice-based machine learning models can detect disease signatures even before overt motor signs appear. Embedding such tools into clinical workflows could enable earlier interventions and enrich our understanding of how PD manifests across neurological and vocal pathways. Voice, long considered secondary in PD diagnostics, may thus become a primary lens for early, scalable, and equitable disease detection.

## Data Availability

The UCI Machine Learning Repository Parkinson’s classification dataset is accessible at https://archive.ics.uci.edu/dataset/174/parkinsons [15, 16].

